# Digital gait measures capture 1-year progression in early-stage spinocerebellar ataxia type 2

**DOI:** 10.1101/2023.10.08.23296692

**Authors:** Jens Seemann, Lina Daghsen, Mathieu Cazier, Jean-Charles Lamy, Marie-Laure Welter, Martin A. Giese, Matthis Synofzik, Alexandra Durr, Winfried Ilg, Giulia Coarelli

**Affiliations:** Section Computational Sensomotorics, Hertie Institute for Clinical Brain Research, Tübingen, Germany; Centre for Integrative Neuroscience (CIN), Tübingen, Germany; Sorbonne Université, Paris Brain Institute - ICM, Inserm, CNRS, AP-HP, Paris; Division Translational Genomics of Neurodegenerative Diseases, Hertie-Institute for Clinical Brain Research and Center of Neurology, University of Tübingen, Tübingen, Germany; German Center for Neurodegenerative Diseases (DZNE), DZNE Tübingen, Germany

**Author notes:** Corresponding authors: Winfried Ilg, Section Computational Sensomotorics, Hertie Institute for Clinical Brain Research, Otfried-Müller-Straße 25, 72076 Tübingen, Germany, Giulia Coarelli, Institut du Cerveau / Paris Brain Institute • ICM, Hôpital de la Pitié-Salpêtrière, CRMR Neurogénétique, Sorbonne Université UM75– Inserm U 1127 – CNRS UMR 7225, 47 boulevard de l’Hôpital, CS21414, 75646, Paris, France. **Disclosures:** J. Seemann, L. Daghsen, M. Cazier, J. Lamy, ML. Welter, A. Giese, and G. Coarelli report no disclosures. Prof. Durr serves as an advisor to Critical Path Ataxia Therapeutics Consortium and her institution (Paris Brain institute) receives her consulting fees from Pfizer, Huntix, UCB, Reata, PTC Therapeutics as well as research grants from the NIH, Biogen, Servier, and the National Clinical Research Program and she holds partly a Patent B 06291873.5 on “Anaplerotic Therapy of Huntington’s Disease and other polyglutamine diseases (2006). Prof. Synofzik has received consultancy honoraria from Ionis, UCB, Prevail, Orphazyme, Servier, Reata, GenOrph, AviadoBio, Biohaven, Zevra, and Lilly, all unrelated to the present manuscript. Dr. Ilg received consultancy honoraria by Ionis Pharmaceuticals, unrelated to the present work.

**Keywords:** gait, spinocerebellar ataxia, SCA2, motor performance measure, longitudinal analysis Statistical analysis was conducted by Winfried Ilg

## Abstract

**BACKGROUND:** With disease-modifying drugs in reach for cerebellar ataxias, fine-grained digital health measures are highly warranted to complement clinical and patient-reported outcome measures in upcoming treatment trials and treatment monitoring. These measures need to demonstrate sensitivity to capture change, in particular in the early stages of the disease.

**OBJECTIVE:** To unravel gait measures sensitive to longitudinal change in the - particularly trial-relevant- early stage of spinocerebellar ataxia type 2 (SCA2).

**METHODS:** Multi-center longitudinal study with combined cross-sectional and 1-year interval longitudinal analysis in early-stage SCA2 participants (n=23, including 9 pre-ataxic expansion carriers; median *ATXN2* CAG repeat expansion 38±2; median SARA [Scale for the Assessment and Rating of Ataxia] score 4.83±4.31). Gait was assessed using three wearable motion sensors during a 2-minute walk, with analyses focusing on gait measures of spatiotemporal variability shown sensitive to ataxia severity, e.g. lateral step deviation.

**RESULTS:** We found significant changes for gait measures between baseline and 1-year follow-up with large effect sizes (lateral step deviation p=0.0001, effect size r_prb_=0.78), whereas the SARA score showed no change (p=0.67). Sample size estimation indicates a required cohort size of n=43 to detect a 50% reduction in natural progression. Test-retest reliability and Minimal Detectable Change analysis confirm the accuracy of detecting 50% of the identified 1-year change.

**CONCLUSIONS:** Gait measures assessed by wearable sensors can capture natural progression in early-stage SCA2 within just one year – in contrast to a clinical ataxia outcome. Lateral step deviation thus represents a promising outcome measure for upcoming multi-centre interventional trials, particularly in the early stages of cerebellar ataxia.

## Introduction

With disease-modifying drugs on the horizon for degenerative ataxias^1-4^, sensitive performance measures are highly warranted. Gait disturbance often presents as a first sign of cerebellar ataxia^5-7^ and is one of the most patient-reported disabling features throughout the disease course^8-10^; thus suggesting a high potential for gait performance measures as both progression and response markers in upcoming treatment trials^3^.

To date, gait measures, including step variability, have demonstrated their sensitivity to ataxia severity mostly in cross-sectional studies of degenerative cerebellar diseases (see reviews in ^6, 11-13^), including also specifically SCA2^14, 15^. However, correlations with clinical scores could be strongly influenced by the range of disease severity^16^. In cohorts spanning a wide range of disease stages, many gait measures – including non-specific ones such as speed – show correlations with disease severity that are often predominantly driven by subjects at both ends of the disease severity spectrum^16^. Notably, in interventional trials, the aim of assessing motor performance measures is qualitatively different: namely to quantify individual change in short trial-like time frames (e.g. one year) – and here often only in a rather specific disease severity stratum

Therefore, to serve as valid markers for capturing change – whether natural history or treatment response change –, gait measures need to demonstrate their sensitivity to individual longitudinal change over those time frames and in those disease severity strata that are relevant for interventional trials ^3, 17^. Here, we present longitudinal gait data from a multi-centre SCA2 cohort collected using wearable sensors in a trial-relevant time frame and disease severity stratum. Specifically, we show that digital measures of motor performance allow to capture longitudinal changes within 1-year in an early-stage SCA2 population where clinical ataxia scores failed to show sensitivity to change.

## Methods

### Patients

Spinocerebellar ataxia type 2 (SCA2) individuals were recruited from the French National Reference Center for Rare Diseases “Neurogenetics” in Paris, Pitié-Salpêtrière Hospital (n=15, assessed 2020-2022) and from the Ataxia Clinic of the University Hospital Tübingen (n=8, assessed 2020-2022). Individuals were included based on the following inclusion criteria: 1.) presence of a CAG repeat in the *AXTN2* gene ≥32; 2.) age: 18-75 years; 3.) SARA score ≤15; 4.) able to walk without walking aids. The exclusion criteria were: severe visual or hearing impairment, cognitive impairment, or orthopaedic limitations. The study population comprised 15 participants at the ataxic stage as defined by a Scale for the Assessment and Rating of Ataxia (SARA)^18^ score of ≥3 (subgroup SCA2_ATX_), and 9 subjects in the pre-ataxic stage (SARA score <3)^18^ (subgroup SCA2_PRE_).

*Estimated time from onset* was defined as the dicerence between current age and estimated age at onset^19^, with estimated disease onset calculated based on the individual’s CAG repeats, as described in^20^. Negative values denote estimated disease onset in the future, positive values denote estimated disease onset in the past.

Healthy controls (n=33) consisted of expansion-negative first-degree relatives of SCA2 carriers and unrelated healthy individuals, all without signs of neurodegenerative disease upon clinical examination. SARA assessments were performed by expert neurologists (Paris: GC, Tübingen: MS).

The study was approved by the local institutional review boards of both participating centres (NCT04288128, IRCB 2018-A02563-52, CPP 19081-60311 for Paris and 598/2011BO1 for Tübingen). Written informed consent was obtained from all study participants before enrolment.

### Gait assessment

Participants performed a 2-minute walk test on quiet, non-public indoor floors in institutional settings by walking back and forth across lines on the floor that were 20 m apart. Participants were instructed to walk at a comfortable and natural pace. Three Opal inertial sensors (APDM, Inc., Portland, US) were attached to both feet, and the posterior trunk at the level of L5 using elastic Velcro straps. Inertial sensor data were collected and wirelessly streamed to a laptop for automatic generation of gait and balance metrics using Mobility Lab software (APDM, Inc., Portland, US). Stride events, as well as spatiotemporal gait parameters from the motion sensors, were extracted using APDM’s Mobility Lab software (Version 2)^21^, which has been shown to provide good-to-excellent accuracy and repeatability^22, 23^. For each detected stride, the following features were extracted: stride length, stride time, lateral step deviation, and foot angles at initial contact. Turning movements and one stride before and after the turns are excluded from the analysis.

From the rich source of possible gait measures, we adopted a hypothesis-driven approach here, selecting only those measures that were considered promising candidate features based on previous studies:

#### Stride length and stride time variability

Measures of spatiotemporal gait variability, such as step length/stride length and step time/stride time variability, have been shown in cross-sectional studies^15, 24-28^ and a longitudinal study in SCA3 ^29^ to be sensitive to ataxia-related gait changes and to be associated with an increased risk of falls^30^. Variability measures were calculated using the coefficient of variation CV=σ/μ, with the standard deviation normalized to the mean^31^. On this basis, stride length CV (StrideL_CV_) and stride time CV (StrideT_CV_) were determined.

#### Lateral step deviation

In a previous cross-sectional study examining ataxic gait characteristics in laboratory and real-life assessments^32^, we identified lateral step deviation (LatStepDev) and a composite measure of *spatial step variability (SPcmp*, combining StrideL_CV_ and LatStepDev), as most sensitive to ataxia severity^32^. In addition, LatStepDev has recently been shown to be sensitive to short-term therapy-induced improvements in SCA27B ^33^. *LatStepDev* was determined based on three consecutive steps by calculating the absolute perpendicular deviation of the midfoot position from the line connecting the first and the third step ^21, 32^. *LatStepDev* was normalized by stride length and averaged over all strides.

#### Toe-out angle variability

Motivated by^15^, we examined an additional feature of variability, namely *toe-out angle*_*SD*_. Toe-out angle was determined as the lateral angle of the foot during the stance phase, relative to the forward motion of the gait cycle^34^. Increased toe-out angle has been shown to be associated with increased stride width, and variability in stride width is associated with dynamic postural instability^15^.

We also included gait speed as a general indicator of functional mobility.

## Statistics

Differences between groups were determined using the non-parametric Kruskal-Wallis test, with post-hoc analysis using the Mann-Whitney U test. Effect sizes were determined using Cliff’s delta^35^. For longitudinal analyses, repeated measures analyses were performed using the non-parametric Friedman test to determine within-group differences between assessments, with post-hoc analysis using a Wilcoxon signed-rank test for pairwise comparisons. Effect sizes for repeated measures were determined by matched-pairs rank biserial correlation ^36^. We report three levels of significance: (i) uncorrected *:p<0.05: (ii) Bonferroni-corrected for multiple comparisons **:p<0.05/n with n=6: number of gait features analysed; (iii) ***:p<0.001. Spearman’s ρ was used to examine the correlation between gait measures and SARA scores. Statistical analysis was performed using MATLAB (version R2020B). Based on the effect size of longitudinal change, a sample size estimation was performed using G*power 3.1^37^ to determine the required cohort size for different levels of reduction of natural progression by a hypothetical intervention. Test–retest reliability of gait measures was calculated using ICC(2, 1) intraclass correlation coefficient ^38, 39^ and calculating the split-half reliability (dividing the walking task into two 1 minute segments). ICC values <0.5, between 0.5 and 0.75, between 0.75 and 0.9, and >0.90 were considered as poor, moderate, good, and excellent reliability, respectively^38^. The ICC is used to determine the minimum detectable change (MDC), which is critical in determining whether a treatment-related slowing of disease progression can be reliably detected or is lost in the measurement noise^40, 41^.

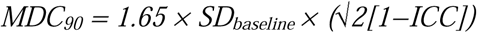

With 1.65 is the z-score of 90 % level of confidence.

## Results

The mean age at baseline assessment was 41.4 ± 12 years [21–66] (SCA2_ATX_: 43.7 ± 9.7 years [28–66]), the mean SARA score was 4.83 ± 6.75 [0–13.5] (SCA2_ATX_: 43.7 ± 9.7 [3.5–13.5]) (Table1+2). In addition, the entire SCA2 population present a mean CAG repeat size of 37.4±2 [32-42], (SCA2_ATX_: 38.2±1.5 [35-42]) and a mean estimated time from onset of 1.78±13.7 years (SCA2_ATX_: 5.34±5.6 years). SARA score was correlated with CAG repeat size (r=0.53, p=0.0088**) and *estimated time from onset* (r=0.61, p=0.0019**) (Table 1+2).

**Table 1.**
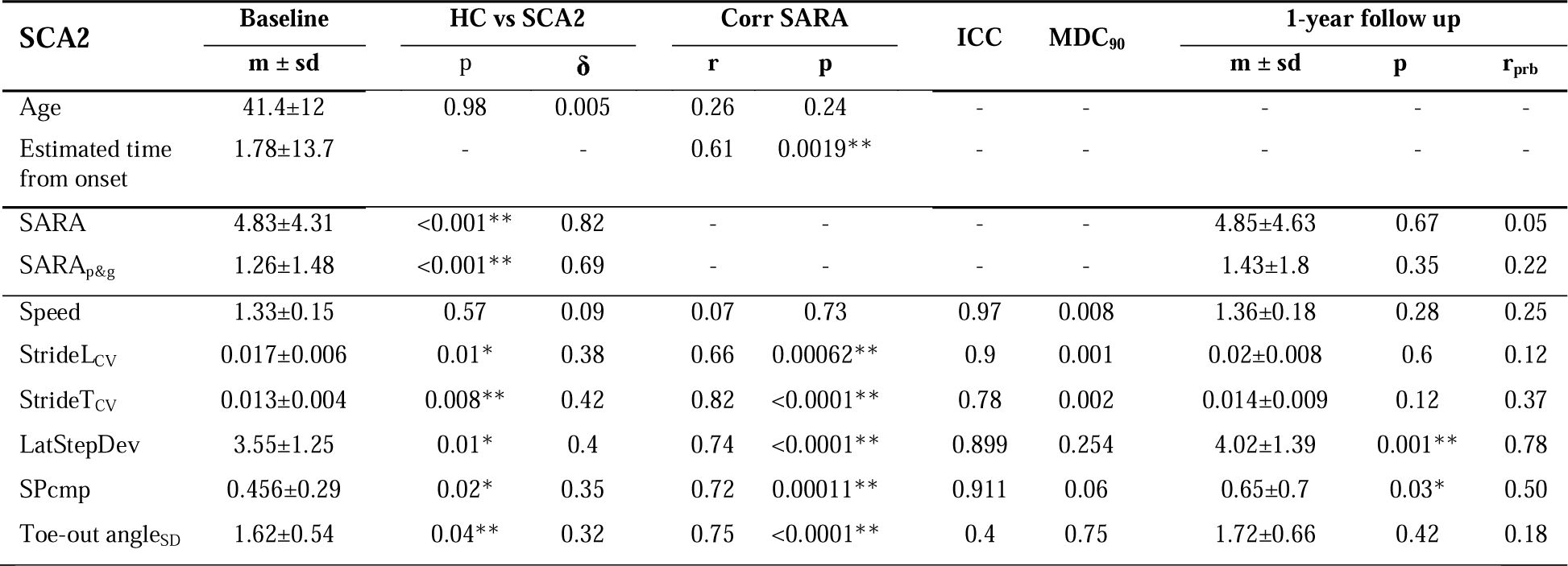
Results of cross-sectional and longitudinal analyses for the SCA2 population. Cross-sectional analyses: Between-group differences in healthy controls (HC) and SCA2 participants for clinical and gait measures. Stars indicate significant between-group differences (*≡ p<0.05, **≡ p<0.0083 Bonferroni-corrected, ***≡ p<0.001). δ indicates the effect size as determined by Cliff’s delta. Correlations between gait measures and clinical ataxia severity (SARA total score, SARA_p&g_ posture&gait subscore) are shown for the SCA2 group. The 3 items of the SARA assessing gait and posture (gait, stance, sitting) were grouped into the SARA posture&gait (SARA_p&g_)subscore ^62, 63^. Effect sizes of correlations are reported using Spearman’s ρ. **Longitudinal analyses of 1-year follow-up assessments:** Paired statistics for within-subject comparisons of clinical scores and gait measures for the two walking conditions (p-values, Wilcoxon signed-rank test; effect sizes r_prb_ determined by matched pairs rank biserial correlation ^36^). m: mean; sd: standard deviation. Analyses are shown for the group of SCA2 subjects at baseline (**SCA2**^**BL**^) and 1-year follow-up (**SCA2**^**FU**^). Estimated time from onset was defined as the di□erence between present age and estimated age at onset^19^, with estimated disease onset calculated based on the individual’s CAG repeats, as described in^20^.

### Cross-sectional analysis of gait measures capturing ataxia severity

Cross-sectional analysis revealed group differences between SCA2 vs HC in all gait measures examined except gait speed (e.g. LatStepDev: p=0.01*, δ=0.4; SPcmp: p=0.02*, δ=0.35; StrideT_CV_: p=0.008**, δ=0.42; Table1). As expected, effect sizes of the group differences became larger for the subpopulation SCA2_ATX_ vs. HC when the pre-ataxic SCA2 mutation carriers were excluded (LatStepDev: δ=0.67; SPcmp: δ=0.61; StrideT_CV_: δ=0.67; see Table 2). No group differences were found between pre-ataxic SCA2 mutation carriers (SCA2_PRE_) and healthy controls for any of the gait parameters (p>0.35).

**Table 2.**
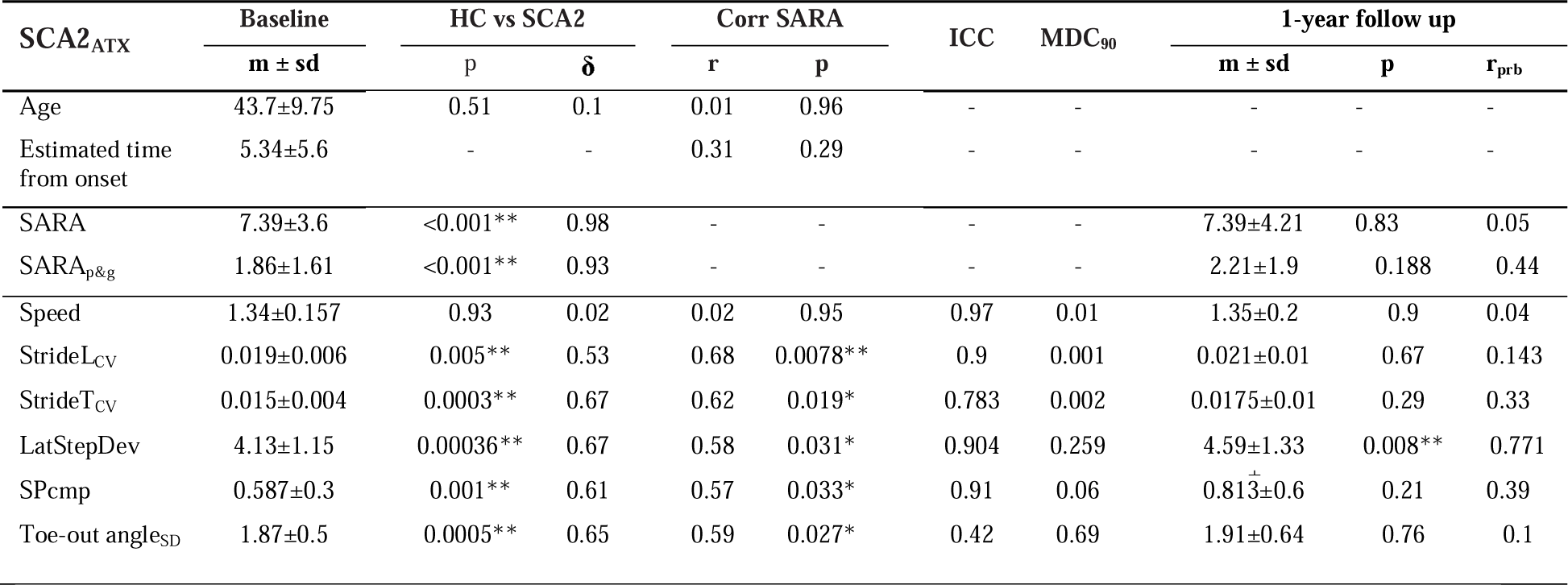
Results of cross-sectional and longitudinal analyses for the SCA2_ATX_ population. Cross-sectional analyses: Differences between groups of healthy controls (HC) and SCA2 subjects for clinical and gait measures. Stars indicate significant between-group differences (*≡ p<0.05, **≡ p<0.0083 Bonferroni corrected, ***≡ p<0.001). δ indicates the effect size as determined by Cliff’s delta. Correlations between gait measures and clinical ataxia severity (SARA total score, SARA_p&g_ posture&gait subscore) are given for the SCA2 group. The 3 items of the SARA assessing gait and posture (gait, stance, sitting) were grouped into the SARA posture&gait (SARA_p&g_) subscore ^62, 63^. Effect sizes of correlations are reported using Spearman’s ρ. **Longitudinal analyses of 1-year follow-up assessments:** Paired statistics for within-subject comparisons of clinical scores and gait measures for the two walking conditions (p-values, Wilcoxon signed-rank test; effect sizes r_prb_ determined by matched pairs rank biserial correlation ^36^). m: mean; sd: standard deviation. Shown are analyses for the group of SCA2 subjects at baseline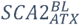and 1-year follow-up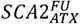. Estimated time from onset was defined as the di□erence between present age and estimated age at onset^19^, with estimated disease onset calculated based on the individual’s CAG repeats, as described in^20^.

Concurrent validity was confirmed for all the ataxia-specific gait measures showing highly significant correlations with the SARA score (e.g. LatStepDev: p<0.0001***, r=0.74; SPcmp: p=0.00011***, r=0.72; StrideT_CV_: p<0.0001***, r=0.82; Figure 1 +Table 1). For these correlation analyses, the effect sizes of the correlations became smaller (but still remained significant) for the subpopulation SCA2_ATX_, due to the smaller range of ataxia severity (Table 2). In addition, gait measures showed correlations to CAG repeat size (LatStepDev: r=0.44, p=0.035*; StrideTCV:r=0.65,p=0.0007**) and *estimated time from onset* (LatStepDev: r=0.44, p=0.037*; StrideTcv: r=0.58, p=0.0041**).

**Figure 1.**
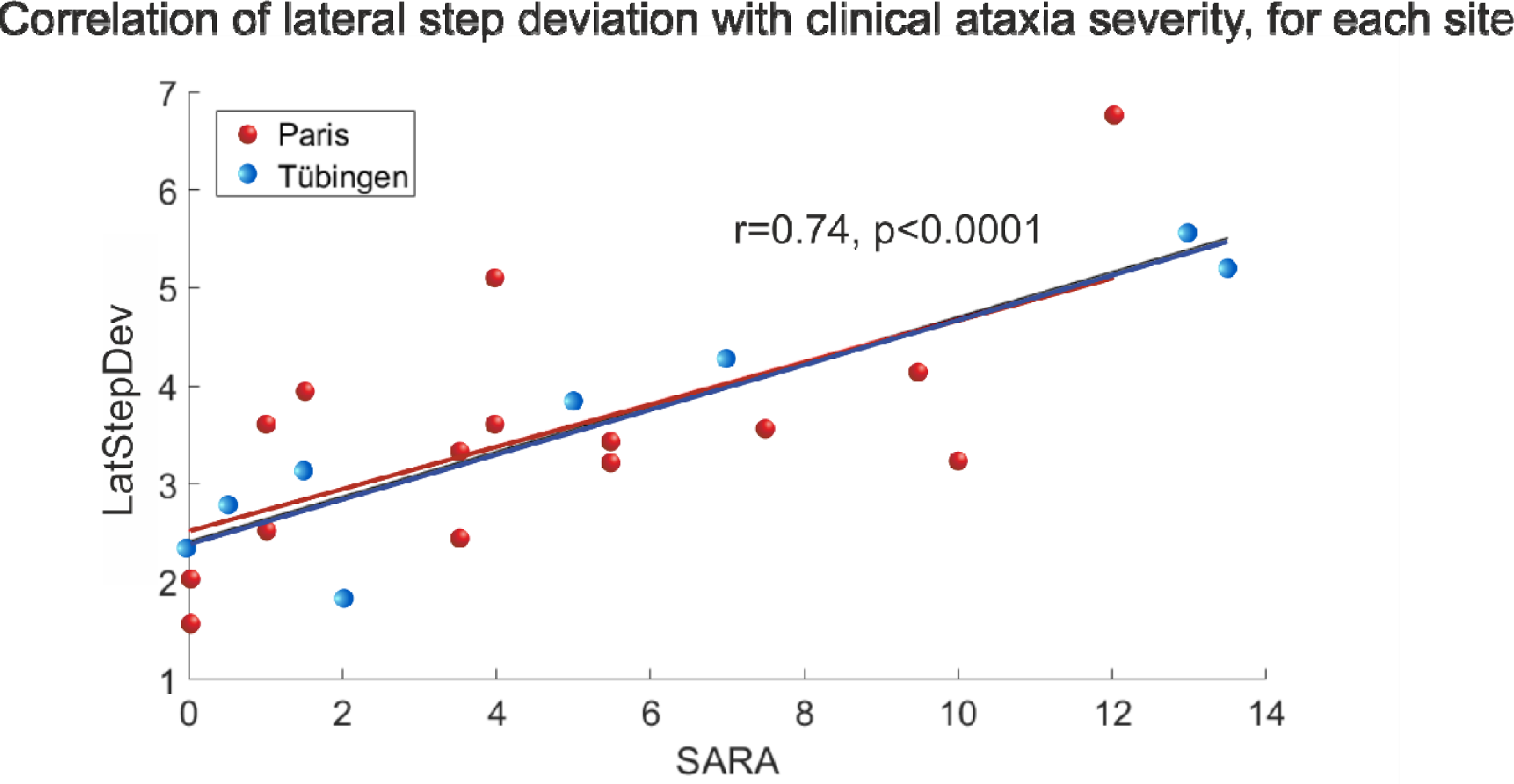
Relationship between the gait measure LatStepDev and the SARA score separately colour coded for participants from both sites, Paris (red) and Tübingen (blue). The lines represent linear fits of the data for each site. Participants from both centers together show a close relationship between LatStepDev and SARA (r=0.74 p<0.0001)***.

### Sensitivity of gait measures to longitudinal change at one year

We next analysed the ability of gait measures to detect longitudinal changes at a 1-year follow-up assessment (time interval: 373±22 days; follow-up data available for all 23 SCA2 participants). While ataxia measured by SARA failed to detect longitudinal change (p=0.67, effect size r_prb_=0.05) (Table1), paired statistics revealed differences between baseline and follow-up for the gait measures LatStepDev (p=0.001**, r_prb_=0.76) and SPcmp (p=0.03*, r_prb_=0.5; Table1, Figure 2A).

**Figure 2.**
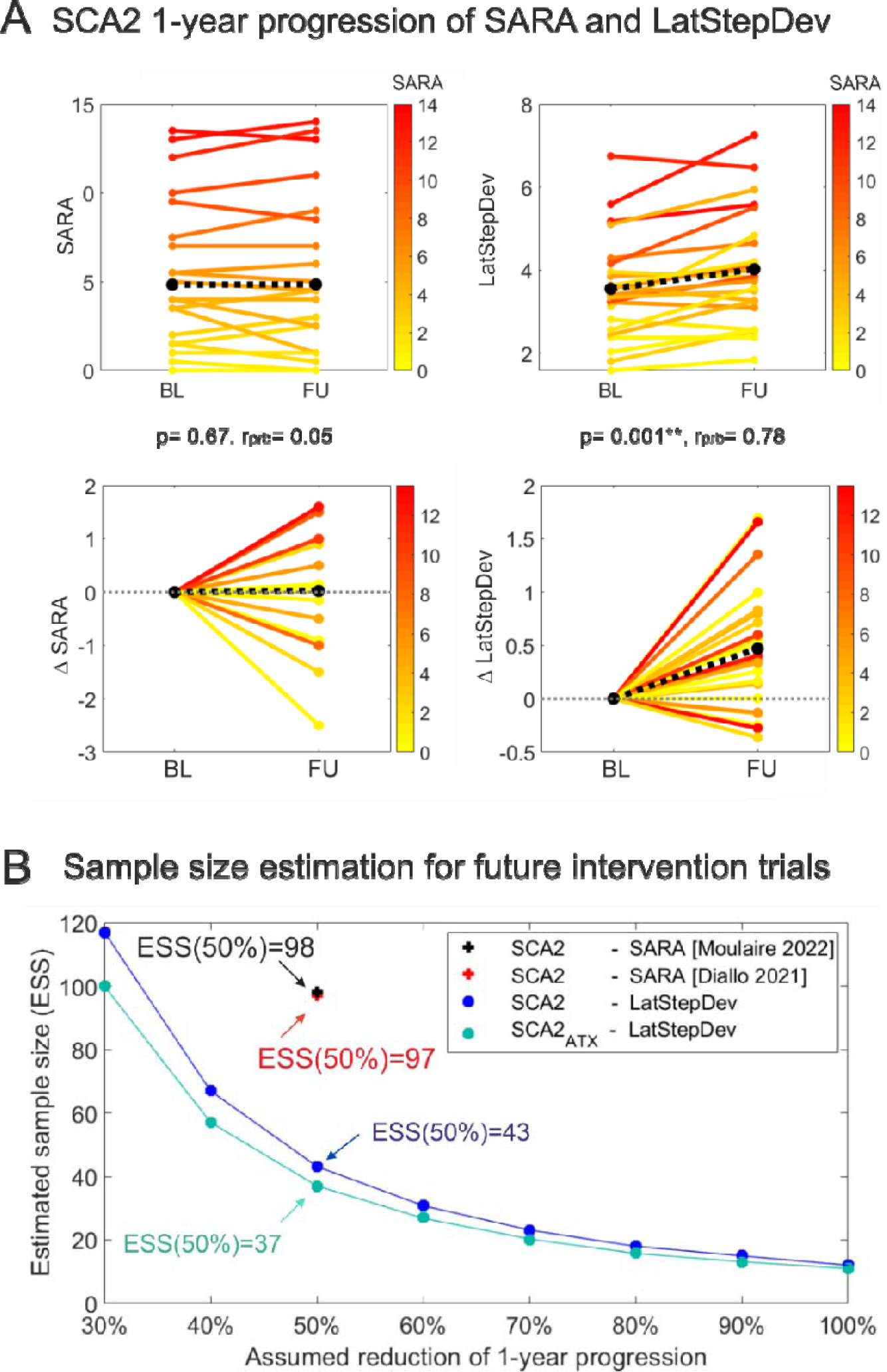
(A) Longitudinal analyses of the 1-year follow-up assessments: Within-subject changes between baseline and 1-year follow-up for the group of SCA2 subjects. Upper panel: Within-subject changes in SARA score and the gait measure LatStepDev at baseline (BL) and 1-year follow-up (FU). Lower panel: Within-subject changes between baseline and 1-year follow-up, expressed as delta (Δ). In all panels, SARA scores of individual cerebellar subjects are colour coded. Black dotted line = mean change across all subjects. Stars indicate significant differences between time points (*≡ p<0.05, **≡ p<0.0083 Bonferroni corrected, ***≡ p<0.001). Effect sizes r_prb_ were determined by matched-pairs rank biserial correlation. (B) Sample size estimates were performed for future intervention trials showing different levels of progression reduction for the gait measure LatStepDev for both the entire SCA2 population and the subpopulation SCA2_ATX_. The estimated number of subjects per study arm is plotted against the hypothesized therapeutic effect for reducing the 1-year progression in SCA2 subjects. Concrete numbers of sample sizes are given to detect a 50% reduction in natural history progression with a hypothetical intervention (80% power and two-sided 5% type I error). For comparison, sample sizes of n=97 (^42^, red cross), and n=98 (^43^, black cross) have recently been reported.

Given the largest effect size, LatStepDev was selected for sample size calculation. To detect a 50% reduction in natural progression with a hypothetical intervention (80% power and two-sided 5% type I error), n=43 subjects would be required using the LatStepDev as the primary outcome measure (Figure 2B). Subgroup analyses revealed an even higher effect size on longitudinal change for the ataxic 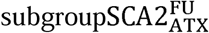 (LatStepDev, r_prb_=0.771) (Table 2), resulting in a reduced estimated sample size of n=37 (Figure 2B). Test-retest reliability and Minimal Detectable Change (MCD) analysis confirm the accuracy of detecting a 50% reduction in identified 1-year change (Table 1).

In contrast, it was not possible to calculate a sample size estimate for the SARA score because it did not show a 1-year change. For comparison, we have included in Figure 2B the sample sizes of recent studies (^42^, n=97, red cross) and (^43^, n=98, black cross) which were reported for SCA2 populations with more advanced disease stages (e.g.: median 10.5 SARA points in^43^) (Figure 2).

## Discussion

Gait disturbance often presents as first sign of cerebellar ataxia^5, 6^ and is one of the most disabling patient-relevant feature throughout the disease course^8-10^, suggesting a high potential as a marker for capturing change-whether related to disease progression or treatment response -in upcoming treatment trials^1, 2, 4, 44^.

This study aimed to test the sensitivity of gait measures to detect ataxia-related longitudinal changes in a time-relevant time frame (one year) and disease severity stratum (early stage) in a SCA2 population in a multicentre setting. Analyses showed that gait measures (i) correlate with cross-sectional clinical ataxia severity, indicating a valid capture of clinical ataxia dysfunction; and in particular (ii) capture longitudinal change between baseline and 1-year follow-up with high effect sizes, substantially outperforming the currently most widely used clinical ataxia scale.

### Gait measures are sensitive to cross-sectional ataxia severity

Our analysis of gait variability measures confirmed the cross-sectional results of previous studies (reviews in^6, 11, 13^), including SCA2 ^14, 15^, which showed (i) a significant difference between healthy controls and ataxia patients and (ii) a high correlation with ataxia severity as measured by the SARA score (Table 1+2).

Importantly, our results validate these findings in a multi-centre setting and in an early-stage SCA2 cohort (SARA score: mean 4.7 points), suggesting their applicability to early disease stages of SCA, which presents the disease severity stratum targeted by upcoming interventional trials^3, 4, 45^.

At the same time, our cross-sectional results illustrate the impact of ataxia severity even within the early-stage study population. When distinguishing between SCA2 patients and healthy subjects, the SCA2_ATX_ subpopulation increased the effect size compared to the overall population SCA2, as pre-ataxic participants SCA2 showed less change in gait measures compared to healthy subjects. (see Tables 1 and 2).

In contrast, the overall SCA2 population shows a larger effect size (compared to SCA2_ATX_) in the correlation of gait measures with the severity of ataxia as measured by the SARA score, due to the wider range of disease stages (namely, including the pre-ataxic stage) (Tables 1 +2).

### Gait measures, not SARA score, capture longitudinal change within one year

In addition to the concurrent validity shown by the correlation with the SARA score, it is crucial for future interventional studies that sensitivity to change is demonstrated by quantifying individual changes in short, trial-like time frames. To date, very few longitudinal gait studies have been conducted in cerebellar ataxia^29, 46-48^, most of them monocentric with heterogeneous populations and gait assessment approaches that are not easily transferable to international multicentre trials. Wearable IMU (inertial measurement unit) sensor technology for quantifying gait has recently become feasible and reliable for large, multicenter clinical trials without sophisticated gait laboratories or expert researchers, making IMUs are easy to use in clinical settings^17^.

Here we now show in a multi-centre setting using wearable motion sensors that the gait measure LatStepDev can quantify these longitudinal changes within a 1-year duration in an early-stage SCA2 population (SARA 4.87±4.28, including 9 pre-ataxic mutation carriers). Effect size was even slightly higher in the subpopulation SCA_ATX_ with manifest ataxia (SARA 7.1±3.6).

In contrast, we did not observe a 1-year longitudinal change in the SARA score in either the entire SCA2 population or the subpopulation SCA2_ATX_. Previous studies in SCA2 that found a longitudinal change in SARA score ^42 49 43^ were performed with more advanced disease stages (e.g.: mean SARA ≥10). These differences can be explained by previous results^50^, reporting an annual delta in SARA of 2.45 points in SCA2 for patients with a disease duration of more than 10 years, but only an average progression of 0.35 SARA points for patients with a disease duration of less than 10 years. This finding again highlights the need to analyse the performance metrics of outcome measures (clinical, digital-motor, etc) in a disease stage-specific fashion.

### Sample size estimates for future trials and Minimal Detectable Change (MDC)

For future disease-modifying drug trials in SCA, the primary goal will be to slow disease progression in a limited trial period, ideally within 1 year^3, 4, 44^. To demonstrate a 50% reduction in natural history with a hypothetical intervention using LatDevStep as the primary outcome measure, n=43 subjects would be required for an early SCA2 population including pre-ataxic mutation carriers, and n=37 for an ataxic SCA2 population including only ataxic mutation carriers. Minimal detectable change (MDC) analysis confirms the accuracy of detecting a 50% reduction in identified 1-year changes (Table 1).

In summary, the large effect sizes and good reliability of this digital-motor measure also in multi-centre settings allow for substantially reduced sample size estimates compared to the SARA for the detection of reduced disease progression within one year (Figure 2). This reduction in sample size could be decisive for the feasibility of a treatment trial: while trials with e.g. 100 SCA2 subjects per trial arm (as required for SARA as outcome) are almost impossible, 37 SCA2 subjects (as required for the gait performance measure LatStepDev in SCA2_ATX_) are well feasible.

### Meaningfulness and ecological validity

To properly evaluate treatment effects in both clinical trials and individual patient treatment settings, it is crucial to identify outcome measures that can detect meaningful changes for patients ^51, 52^. Gait assessment can provide meaningful outcome measures for evaluating treatment interventions, as cerebellar ataxia patients report gait and functional mobility impairments as having the greatest impact on their daily lives^8-10^. While longitudinal studies relating differences in gait measures to patient-reported outcomes are still lacking, we have shown in^32^ that the gait performance measure LatStepDev is highly correlated with the patient-reported subjective balance confidence (ABC score^53^). In particular, LatStepDev has been shown to capture ataxia-related gait impairments in real-life walking behavior, the latter being particularly important for demonstrating ecological relevance ^32, 54, 55^. In addition, LatStepDev has recently been shown to be sensitive to short-term therapy-induced improvements in SCA27B^33^ in correspondence with a change in a key patient reported outcome (Patient Global Impression, PGI) ^52^.

### Study limitations

Our findings are limited by the relatively small cohort size. In particular, our study cohort was not sufficiently powered for detecting longitudinal change within the pre-ataxic group only. Thus, larger future studies, including a larger number of pre-ataxic subjects, are needed to further validate the promises of gait measures and relate longitudinal changes in gait to patient-centered outcomes and patient-meaningful aspects of health^52, 56^ as well as to corresponding changes in molecular (such as blood neurofilament light chain ^57, 58^) and imaging biomarkers^59^.

## Conclusion

Our study demonstrates that digital gait measures allow to capture natural history progression change of SCA2 within one year, with effect sizes exceeding the main clinical rating scale (SARA) -which is still the most widely established outcome measure in this field. The proposed gait measures can be reliably captured by wearable motion sensors in multi-centre studies including centres without sophisticated motion laboratories and expert researchers. In particular the digital gait measure LatStepDev represents a promising performance outcome for future SCA intervention trials, particularly in the early stages of the disease, which are also more representative of the disease strata that will be enrolled in future trials than the advanced stages of the disease ^4, 45, 60, 61^

## Data Availability

Data will be made available upon reasonable request. The authors confirm that the data supporting the findings of this study are available within the article. Raw data regarding human subjects (e.g. clinical data) are not shared freely to protect the privacy of the human subjects involved in this study; no consent for open sharing has been obtained.

## Acknowledgments

We would like to thank all the participants including in this study. We would like to thank BIOGEN and IONIS which funded the NCT04288128 study and INSERM, which sponsored the NCT04288128 study (to A. D.). This work was supported by the International Max Planck Research School for Intelligent Systems (IMPRS-IS) (to J.S.) and the Else Kröner-Fresenius-Stiftung Medical Scientist programme ‘ClinbrAIn’ (to W.I.). as well as the Else Kröner-Fresenius Stiftung Clinician Scientist programme “PRECISE.net” (to M.S.). Work on this project was supported, in part, by the Deutsche Forschungsgemeinschaft (DFG, German Research Foundation) No 441409627, as part of the PROSPAX consortium under the frame of EJP RD, the European Joint Programme on Rare Diseases, under the EJP RD COFUND-EJP N° 825575 (to M.S. and A.D.).

## Author Roles

1. Research project: A. Conception, B. Organization, C. Execution;
2. Statistical Analysis: A. Design, B. Execution, C. Review and Critique;
3. Manuscript Preparation: A. Writing of the first draft, B. Review and Critique;

W.I.: 1A, 1B, 2A, 3A

J.S.: 1C, 2C, 3B

M.S.: 1A, 2C, 3A

G.C.: 1B, 1C, 3B

A.D.: 1B, 2C, 3B

L.D.: 1B, 1C, 3B

M.C.: 1B, 1C, 3B

J.C.L.: 1B, 1C, 3B

M.L.W.: 1B, 1C, 3B

M.G.: 1B, 1C, 3B

